# Is Blood Type Associated with COVID-19 Severity?

**DOI:** 10.1101/2020.08.11.20172676

**Authors:** Angelico Mendy, Jason L. Keller, Senu Apewokin, Ardythe L. Morrow

**Affiliations:** Division of Epidemiology, Department of Environmental and Public Health Sciences, University of Cincinnati College of Medicine, Cincinnati, OH, USA; Center for Health Informatics, Department of Biomedical Informatics, College of Medicine, University of Cincinnati, OH USA; Division of Infectious Diseases, Department of Medicine, University of Cincinnati College of Medicine, Cincinnati, OH, USA

**Author notes:** **Corresponding Author**: Angelico Mendy, MD, PhD, Division of Epidemiology, Department of Environmental & Public Health Sciences, The University of Cincinnati College of Medicine, 160 Panzeca Way, Room 335, Cincinnati, OH 45267.

**Keywords:** Blood Type, ABO, Rhesus, COVID-19, SARS-CoV-2

## Abstract

Blood type purportedly influences susceptibility to severe acute respiratory syndrome coronavirus-2 (SARS-CoV-2) infection, but whether it affects severity of coronavirus disease 2019 (COVID-19) is unclear. Therefore, we examined the association of blood type and rhesus with hospitalization and disease severity among 428 COVID-19 patients diagnosed at the University of Cincinnati health system. In the sample, 50.2% of participants had the blood type O, 38.7% had the blood type A, 17.5% had the blood type B, and 3.5% had the blood type AB. In analysis adjusted for sociodemographic characteristics and comorbidities, the blood types A (OR: 0.90, 95% CI: 0.54, 1.50), B (OR: 0.93, 95% CI: 0.51, 1.69), AB (OR: 0.69, 95% CI: 0.20, 2.41), and O (OR: 1.18, 95%: 0.74, 1.98) were not associated with hospitalization for COVID-19. Similarly, the blood types A (OR: 0.93, 95% CI: 0.52, 1.65), B (OR: 0.92, 95% CI: 0.46, 1.84), AB (OR: 0.30, 95% CI: 0.04, 2.44), and O (OR: 1.25, 95%: 0.73, 2.14) were not associated with admission to intensive care unit or death in COVID-19. In conclusion, blood type is not associated with hospitalization or disease severity in COVID-19; therefore, it may not be useful marker for identifying patients at risk for severe COVID-19.

---

Since its occurrence in China in December 2019, the coronavirus disease 2019 (COVID19) has spread into a pandemic by March 2020 to become a major global public health crisis.^1^ The disease is caused by the recently identified severe acute respiratory syndrome coronavirus-2 (SARS-CoV-2), a highly contagious virus transmitted through droplets, contaminated surfaces, and aerosols within closed places.^2,3^ Although COVID-19 is asymptomatic in some individuals, in others it can cause manifestations ranging from mild disease with dry cough and dyspnea to severe pneumonia requiring admission to intensive care unit and sometimes leading to death.^3^

The susceptibility to SARS-CoV-2 infection has recently been suggested to be influenced by blood type, with type A reportedly being associated with increased risk, whereas type O could protect against the infection.^4,5^ This may be due to the ability of anti-A antibodies to prevent cellular entry of SARS-CoV-2 by inhibiting the binding between the spike (S) viral protein and cellular angiotensin converting enzyme 2 (ACE-2) receptors.^6^ Moreover, *in vitro* studies propose that blood type might affect von Willebrand factor and factor VIII to modify the risk of thrombosis, a common complication of COVID-19.^7^ Yet, epidemiologic studies on blood type and COVID-19 severity among SARS-CoV-2 infected patients are lacking; therefore, we aimed to examine the association of blood type with severe COVID-19 in a racially and ethnically diverse cohort.

## METHODS

We identified COVID-19 patients diagnosed between March 13, 2020 and July 5, 2020 in the electronic medical record system of the University of Cincinnati health system which consists of hospitals and clinics located in the states of Ohio, Kentucky, and Indiana. COVID-19 diagnosed using nasopharyngeal reverse transcriptase polymerase chain reaction test for SARS-CoV-2. Our study included all COVID-19 patients who had complete data on ABO blood and the studied outcomes such as hospitalization defined as admission to the hospital for at least 24 consecutive hours as well as severe disease defined as admission to ICU and/or death.^3^

Date of birth, sex, race/ethnicity, and smoking were self-reported. Comorbidities were defined using the 10^th^ revision of the International Classification of Diseases (ICD10) and included obesity (ICD10 code E66), diabetes (ICD10 codes E10 and E11), asthma (ICD10 code J45), chronic obstructive pulmonary disease (COPD) (ICD10 code J44), cardiovascular disease (CVD) (ICD10 code I00-I99), chronic kidney disease (CKD) (ICD10 code N18), and history of cancer (ICD10 codes C00-D49).^3^

Descriptive analyses were performed to summarize the characteristics of study participants overall and by ABO blood type. The odds ratios (OR) and corresponding 95% confidence intervals (CI) for the associations of ABO blood type with hospitalization and disease severity were estimated by means of logistic regression analysis. The models were adjusted for age, sex, race/ethnicity, and cigarette smoking as well as the comorbidities (obesity, diabetes, asthma, COPD, CVD, CKD, and history of cancer). The analyses were performed in SAS Version 9.4 (SAS Institute, Cary, NC) with two-sided *P*-values < 0.05 considered statistically significant.

## RESULTS

A total of 428 COVID-19 patients were included in the study. They had a median age of 45.4 years (interquartile range: 31.2, 63.5) and 37.4% of them were men. The majority of participants were Non-Hispanic Black (53.0%); 21.5% were Hispanic, 21.0% were non-Hispanic White, and 4.4% were of ‘Other’ race/ethnicity. The comorbidities reported in the patients included CVD (64.0%), obesity (34.3%), diabetes (33.9%), history of cancer (30.1%), asthma (18.7%), and COPD (14.5%). Regarding COVID-19 outcomes, 192 patients (44.9%) were hospitalized and 101 (23.6%) had a severe form of the disease (Table 1).

**Table 1:** Characteristics of study participants overall and by blood type (N = 428)

| Characteristics | All | Blood Type A |  |  | Blood Type B |  |  | Blood Type AB |  |  | Blood Type O |  |  |
| --- | --- | --- | --- | --- | --- | --- | --- | --- | --- | --- | --- | --- | --- |
|  |  | Non-A | A | P-value | Non-B | B | P-value | Non-AB | AB | P-value | Non-O | O | P-value |
| Prevalence, % | 100 | 71.3 | 28.7 |  | 82.5 | 17.5 |  | 96.5 | 3.5 |  | 49.8 | 50.2 |  |
| **Socio-demographics** |  |  |  |  |  |  |  |  |  |  |  |  |  |
| Age, years, median (SE) | 45.4 (1.7) | **40.8 (1.5)** | **50.5 (2.8)** | **0.044** | 44.4 (1.8) | 49.5 (3.3) | 0.080 | 45.4 (1.8) | 44.0 (8.7) | 0.927 | **49.7 (2.0)** | **38.9 (1.3)** | **0.001** |
| Male sex, % | 37.4 | 37.4 | 37.4 | 0.997 | 36.3 | 42.7 | 0.298 | 37.5 | 33.3 | 0.741 | 39.0 | 35.8 | 0.500 |
| Race/ethnicity, % |  |  |  | **< 0.001** |  |  | **< 0.001** |  |  | **0.030** |  |  | **< 0.001** |
| Non-Hispanic Whites | 21.0 | **16.4** | **32.5** |  | **23.8** | **8.0** |  | **20.6** | **33.3** |  | **23.9** | **18.1** |  |
| Non-Hispanic Blacks | 53.0 | **53.8** | **32.5** |  | **47.3** | **80.0** |  | **53.0** | **52.3** |  | **61.5** | **44.7** |  |
| Hispanics | 21.5 | **26.6** | **8.9** |  | **24.6** | **6.7** |  | **22.3** | **0** |  | **7.5** | **35.3** |  |
| Other | 4.4 | **3.3** | **7.3** |  | **4.2** | **8.0** |  | **4.1** | **13.3** |  | **7.0** | **1.9** |  |
| Past & current smokers, % | 39.7 | 37.4 | 45.5 | 0.155 | 37.4 | 50.7 | 0.096 | 40.4 | 20.0 | 0.231 | **45.5** | **34.0** | **0.014** |
| **Comorbidities** |  |  |  |  |  |  |  |  |  |  |  |  |  |
| Obesity, % | 34.3 | 32.8 | 38.2 | 0.285 | 33.7 | 37.3 | 0.548 | 34.9 | 20.0 | 0.234 | 36.6 | 32.1 | 0.324 |
| Diabetes, % | 33.9 | 31.5 | 39.8 | 0.098 | 33.1 | 37.3 | 0.486 | 33.7 | 40.0 | 0.610 | **39.0** | **28.8** | **0.027** |
| Asthma, % | 18.7 | 17.7 | 22.0 | 0.272 | 17.6 | 24.0 | 0.194 | 18.6 | 20.0 | 1.00 | **22.5** | **14.9** | **0.042** |
| COPD, % | 14.5 | 13.8 | 16.3 | 0.508 | 13.3 | 20.0 | 0.135 | 14.5 | 13.3 | 1.00 | 17.4 | 11.6 | 0.091 |
| CVD, % | 64.0 | **61.0** | **71.5** | **0.039** | 62.6 | 70.7 | 0.186 | 63.9 | 66.7 | 0.828 | **70.9** | **57.2** | **0.003** |
| CKD, % | 22.4 | 20.0 | 28.5 | 0.058 | 21.0 | 29.3 | 0.114 | 22.5 | 20.0 | 1.00 | **28.2** | **16.7** | **0.005** |
| History of cancer, % | 30.1 | 29.2 | 32.5 | 0.496 | 28.6 | 37.3 | 0.135 | 30.0 | 33.3 | 0.784 | 34.3 | 26.0 | 0.064 |
| **COVID-19 Outcomes** |  |  |  |  |  |  |  |  |  |  |  |  |  |
| Hospitalization, % | 44.9 | 44.6 | 45.5 | 0.860 | 44.8 | 45.3 | 0.928 | 45.3 | 33.3 | 0.361 | 44.6 | 45.1 | 0.915 |
| Severe COVID-19, % | 23.6 | 23.9 | 22.8 | 0.796 | 23.5 | 24.0 | 0.928 | 24.2 | 6.7 | 0.211 | 22.1 | 25.1 | 0.457 |
| Admission to ICU, % | 21.3 | 21.6 | 20.3 | 0.764 | 21.5 | 20.0 | 0.769 | 21.8 | 6.7 | 0.210 | 19.2 | 23.3 | 0.311 |
| Death, % | 7.5 | 6.2 | 10.6 | 0.122 | 7.6 | 6.7 | 0.769 | 7.7 | 0.0 | 0.616 | 8.5 | 6.5 | 0.446 |

Abbreviations: SE: standard error; COPD: chronic obstructive pulmonary disease; CVD: cardiovascular disease; CKD: chronic kidney disease; ICU: intensive care unit. P-values for differences in age calculated using Wilcoxon rank tests and P-values for difference in prevalence of characteristics calculated using chi-square or Fisher's exact test.

About half of the study participants were of the blood type O (50.2%); 38.7% had the blood type A; 17.5% had the blood type B, and 3.5% had the blood type AB. Participants with the blood type A tended to be older, to be non-Hispanic White, and to have more CVD compared to those with non-A blood type. Participants with the blood type B tended to be non-Hispanic Black compared to those with non-B blood type. Participants with the blood type AB tended to be non-Hispanic White compared to those with non-AB blood type. Participants with the blood type O tended to be younger, and to be Hispanic; but they were less likely to have diabetes, asthma, CVD, and CKD compared to those with non-O blood type (Table 1).

In analysis adjusted for sociodemographic characteristics and comorbidities, the blood types A (OR: 0.90, 95% CI: 0.54, 1.50), B (OR: 0.93, 95% CI: 0.51, 1.69), AB (OR: 0.69, 95% CI: 0.20, 2.41), and O (OR: 1.18, 95%: 0.74, 1.98) were not associated with hospitalization. Likewise, the blood types A (OR: 0.93, 95% CI: 0.52, 1.65), B (OR: 0.92, 95% CI: 0.46, 1.84), AB (OR: 0.30, 95% CI: 0.04, 2.44), and O (OR: 1.25, 95%: 0.73, 2.14) were not associated with disease severity in COVID-19 (Table 2). The associations of ABO blood type with hospitalization and severe COVID-19 by rhesus were reported in the Table 2.

**Table 2:** Association of ABO blood type with hospitalization and disease severity in COVID-19

| ABO Blood type | Hospitalization |  |  | Severe COVID-19 |  |  |
| --- | --- | --- | --- | --- | --- | --- |
|  | n/N | OR (95% CI) | P-value | n/N | OR (95% CI) | P-value |
| **Blood type A** |  |  |  |  |  |  |
| Non-A | 136/305 | Reference |  | 73/305 | Reference |  |
| A | 56/123 | 0.90 (0.54, 1.50) | 0.70 | 28/123 | 0.93 (0.52, 1.65) | 0.80 |
| A - | 5/13 | 1.07 (0.29, 3.95) | 0.86 | 2/13 | N/A |  |
| A + | 51/110 | 0.90 (0.53, 1.53) | 0.74 | 26/110 | 0.94 (0.52, 1.70) | 0.88 |
| **Blood type B** |  |  |  |  |  |  |
| Non-B | 158/353 | Reference |  | 83/353 | Reference |  |
| B | 34/75 | 0.93 (0.51, 1.69) | 0.81 | 18/75 | 0.92 (0.46, 1.84) | 0.82 |
| B - | 2/5 | N/A |  | 2/5 | N/A |  |
| B + | 32/72 | 0.97 (0.52, 1.80) | 0.83 | 16/72 | 0.88 (0.43, 1.80) | 0.19 |
| **Blood type AB** |  |  |  |  |  |  |
| Non-AB | 187/413 | Reference |  | 100/413 | Reference |  |
| AB | 5/15 | 0.69 (0.20, 2.41) | 0.56 | 1/15 | 0.30 (0.04, 2.44) | 0.26 |
| AB - | 0/0 | N/A |  | 0 | N/A |  |
| AB + | 5/15 | 0.69 (0.20, 2.41) | 0.56 | 1/15 | 0.30 (0.04, 2.44) | 0.26 |
| **Blood type O** |  |  |  |  |  |  |
| Non-O | 95/213 | Reference |  | 47/213 | Reference |  |
| O | 97/215 | 1.18 (0.74, 1.89) | 0.49 | 54/215 | 1.25 (0.73, 2.14) | 0.42 |
| O - | 7/12 | 1.39 (0.36, 5.30) | 0.72 | 3/12 | 1.02 (0.23, 4.46) | 0.89 |
| O + | 90/203 | 1.20 (0.74, 1.94) | 0.97 | 51/203 | 1.29 (0.74, 2.22) | 0.57 |

Abbreviations: OR: odds ratio; CI: confidence interval.
Odds ratios calculated using logistic regression. Models adjusted for sociodemographic characteristics (age, sex, race/ethnicity, and smoking) and comorbidities (obesity, diabetes, asthma, COPD, CVD, CKD, and history of cancer).

## DISCUSSION

In this study, ABO blood type was not associated with hospitalization or disease severity. Others have reported that individuals with type O blood may be at lower risk of SARS-CoV-2 infection and those with type A could be at increased risk,^4,5^ COVID-19 patients with type O blood were overrepresented in our study sample (50.3%). Patients with type A blood were only 28.7% of our sample. Studies conducted in Chinese and European population had significantly fewer patients with blood type O and more patients with blood type A.^8-10^ However, a US cohort with a racially and ethnically diverse population reported a distribution in blood groups similar to our study.^11^

Our study is one a few on blood group and COVID-19 severity. Consistent with our findings, three studies performed in Turkey and in the US found no association between blood type and disease severity in COVID-19 defined as intubation and/or death.^9,11,12^ Our results are in contradiction however with the study by Zhao et al. who noted that risk of death in COVID-19 patients was increased with blood type A and reduced with blood type O.^4^ An ecological study compared COVID-19 morbidity and mortality of 86 Asian, European, African and American countries with the World distribution of ABO and Rhesus blood groups and found a significant correlation.^13^ However, the study did not include patient-level data and suffered from ecological fallacy, which is the assumption that factors associated with the national rates of a health outcome are associated with the outcome in individual patients.^14^ It has been suggested that ABO blood group are also expressed endothelial cells and platelets and might influence the biological activity of von Willebrand factor and factor VIII. This would result in in a risk thrombosis increased in patients with blood type A and reduced in those with blood type O.^7^

In conclusion, we did not observe an association between blood type and disease severity in COVID-19 patients. Therefore, blood type may not be a useful marker to identify COVID-19 patients at higher risk of severe disease.

## Data Availability

The SAS codes and datasets are available for reproducing the results upon reasonable request

## Funding

Angelico Mendy’s contribution was partly funded by grant P30 ES006096 from the U.S. NIH. The project described was supported by the National Center for Advancing Translational Sciences of the NIH, under Award Number 5UL1TR001425-03. The content is solely the responsibility of the authors and does not necessarily represent the official views of the NIH.

## Disclosures

Authors declare no relationship with industry or other relevant entities that might pose a conflict of interest in connection with the submitted article.

## Author Contributions

AM contributed to the study concept and design, analysis and interpretation of data, and writing of the manuscript. SA and ALM contributed to the interpretation of data and writing of the manuscript. JLK contributed to the collection of the dataset. All authors reviewed the manuscript for intellectual content. AM takes full responsibility for the integrity of the dataset and the analysis results.

## Notes

### Competing Interest Statement

The authors have declared no competing interest.

### Clinical Trial

Not Applicable

### Funding Statement

Angelico Mendy contribution was partly funded by grant P30 ES006096 from the US NIH. The project described was supported by the National Center for Advancing Translational Sciences of the NIH, under Award Number 5UL1TR001425-03.

### Author Declarations

University of Cincinnati Institutional Review Board.

